# “It’s like the spirit of the blitz”: the impact of the COVID-19 pandemic on LTCF staff working relationships in England

**DOI:** 10.1101/2025.06.09.25328141

**Authors:** Danni Collingridge Moore, Natalie Cotterell

## Abstract

**Background:** The COVID-19 pandemic created novel challenges for staff working in the adult social care sector, however its impact on staff working relationships within facilities is unclear. The aim of this study is to explore the effect the pandemic had on staff working relationships.

**Method:** Secondary analysis of data collected in semi-structured, qualitative interviews with LTCF staff. Twenty-four participants working in eight LTCFs in England were recruited to discuss their experiences of working in a LTCF during the COVID-19 pandemic. Thematic analysis was used to explore the impact of the pandemic on staff working relationships.

**Results:** Seven themes were identified, these were a) a shared commitment to providing care to residents, b) strengthening working relationships between staff members, c) adapting to novel and changing roles, d) working as an incentive for socialisation, e) leadership by example, f) recruitment and retraining of new staff, and g) divisions between furloughed and attending LTCF staff.

**Discussion:** The findings show that within the challenges that the pandemic created for LTCFs, staff members reported improved staff working relationships, characteristics by strengthened bonds between staff, improvements in team working and developing new approaches to resident care. However, furloughed and newly recruited staff members were, at times, excluded from these developments. Further research is needed to explore how improvements to staff working relationships can be replicated and sustained prior to future pandemics, and the supportive effect these may have on quality of care and staff mental health and wellbeing.

## Introduction

In the COVID-19 pandemic, one of the worst affected sectors within society was adult social care, impacting service users, their family members and care providers. Since the onset of the pandemic in March 2020, there were 274,063 deaths of long-term care facility (LTCF) residents, 45,632 of which involved COVID-19, accounting for 16.7% of all deaths (1). The health implications that the pandemic brought to this sector were not limited to residents; those employed in caring professions experienced significantly higher mortality rates in terms of deaths involving COVID-19 compared to those of the same age in the general population, with care workers and home carers accounting for the majority of deaths within this group (74.0%) (2).

The UK government issued numerous policies aimed to protect and support LTCF staff during the COVID-19 pandemic (3). Adult social care staff were assigned ‘key worker’ status, and for those who were unable to work due to suspected or confirmed COVID-19 infection, funding was provided to backfill wages, alongside increases to statutory sick pay (4, 5). A national recruitment campaign was developed to attract 20,000 people into adult social care, specifically targeting workers previously recruited from sectors that were unable to operate due to COVID-19 restrictions, such as hospitality or leisure services (6). Funding streams were developed to accommodate the costs associated with staff shortages that resulted from restrictions on working across multiple facilities (7). LTCF managers were supported in identifying, assessing, and developing plans to protect vulnerable workers through risk assessments, allowing staff who were classed as either clinically vulnerable or at a higher risk of infection to be reallocated to roles that did not involve providing direct care to symptomatic residents (8). These strategies were not limited to the UK, with many countries introducing some form of measures that related to recruiting volunteers and workers to the long-term care sector, increasing the maximum number of working hours available to staff, extending visas for foreign workers within the sector, or implementing isolation measures for staff with suspected or confirmed COVID-19 (9).

Despite these policies, the long-term care sector still experienced significant challenges in terms of maintaining and supporting staff (10). In the first months of the pandemic, between mid-April to mid-May, absence rates among LTCF staff were around 10% (11). During this time, the experience of many staff members working in LTCFs was characterised by anxiety and fear, with concerns over limited access to personal protective equipment, difficulties enacting social distancing measures, and delays in testing kits widely reported (8, 12).

The COVID-19 pandemic arguably exacerbated ongoing issues within the long-term care system in England, some of which still exist today. Rates of staff turnover are high; in 2022/2023, 28.3%, or approximately 390,000 people were leaving their posts over the course of the year (although some may have remained in the adult social care sector) (13). Salaries remain relatively low compared to the wider population; the median hourly rate for LTCF workers is £10.11 (61 pence higher than the national living wage), and opportunities to pursue training and education vary between care providers; 54% of workers in this sector have no relevant social care qualifications (13). In an international review of key COVID-19 policies in the long-term care sector using data from Australia, Canada, Spain and the United States, severe staffing shortfalls, skill mix deficits and unmet educational needs were identified as key contributors to the challenges faced, all of which existed in some form prior to the pandemic (14).

The challenges faced by LTCF staff during this time have been explored in previous research studies and independent reports (15-17), however, there has been fewer studies on the impact of the pandemic on the working relationships between LTCF staff during and after the pandemic. Insights into these experiences could be used to inform how to strengthen and support the long-term care workforce and guide the development and implementation of policies in future pandemic.

### Aims and objectives

The aim of this study is to explore the effect of the COVID-19 pandemic on staff working relationships within LTCFs in England.

## Methods

### Study design

The data analysed in this study were collected as part of case studies conducted in eight LTCFs in north-west England. The objective of the original study was to explore the implementation of government issued polices by LTCF staff during the pandemic, and the relative ease and effectiveness of putting these policies into practice with the facility. Further details of the methods have been published in detail elsewhere (18).

### LTCF and participant recruitment

Twenty-four staff members based across eight LTCFs in north-west England were interviewed between October 2023 and April 2024. Information about the study and what taking part would involve was distributed to LTCFs in the region through the Enabling Research in Care Homes Network, who had an existing database of LTCFs who had registered an interest in taking part in research. Inclusion criteria included registration with the Care Quality Commission as of May 2023, registered as providing care to older adults aged 65 years and above, and active during the COVID-19 pandemic.

Staff members who worked in LTCFs that had consented to take part in the study were provided with information packs handed out by the facility manager. Staff were eligible to take part in the study if they were aged 18 years or over, were able to participate in the interview in English, and worked at the facility during the COVID-19 pandemic. Prior to the interview, the research team explained the aims of the study, its funding source, and the background of the researchers. All staff members who participated in an interview provided written informed consent prior to taking part.

### Data collection

Qualitative, semi-structured Interviews were conducted with staff members at a time which was convenient to them. Interviews were either online or in person at the LTCF. Each interview lasted 40–60⍰minutes and were conducted by NC, with DCM observing a subset of the sample. NC and DCM were both employed on the study and have previous training on qualitative research methodology and data analysis. The interview topic guide was developed within the research team to allow LTCF staff members to reflect on their role in implementing the key policy recommendations issued by the UK government for LTCFs during the COVID-19 pandemic. The interviews were audio recorded and transcribed using Microsoft Teams, and the interview transcripts and the associated data was managed, organised and stored using Atlas.ti. software (19). Ethical approval was granted by Lancaster University Faculty of Health and Medicine Research Ethics Committee (reference: FHM-2023-3368-RECR-3).

### Data analysis and reporting

The research team familiarised themselves with the interview transcripts by reading each one. Next, the transcripts were analysed using thematic analysis, guided by the steps developed by Braun and Clarke; familiarisation with the data, generating initial codes, searching for themes, reviewing themes, and finally defining and naming the themes (20). The reporting of the data and study methodology was guided by the ‘Consolidated criteria for reporting qualitative research (COREQ)’, a 32-item checklist to guide the reporting of qualitative research studies (21).

## Findings

Eight LTFCs were recruited; four providing residential care and four providing nursing care. Seven were privately owned and one was not for profit, and number of beds ranged from 25 to 120. Semi-structured interviews were conducted with 24 participants, the majority of whom were female (n=21), 79.2% worked full-time, with ages ranging from 23 to 68 years old (mean-46). In total, five registered LTCF managers; four senior care assistants; three clinical managers; three care assistants; three housekeepers; two Care Home Assistant Practitioners (CHAPs); two deputy care managers; one activity coordinator; and one head of operations were interviewed. Within the sample, 21 identified as White British, two as Indian/British Indian, and one as Portuguese. English was the first language of all but two interviewees. Participants had spent on average 21 years working in direct patient care was (range 5-46) and 18 years (range 5-46) working in LTCFs.

Seven themes were identified in the data (see Table 1 for an overview). These were a) a shared commitment to providing care to residents, b) strengthening working relationships between staff members, c) adapting to novel and changing roles, d) working as an incentive for socialisation, e) leadership by example, f) recruitment and retraining of new staff, and g) divisions between furloughed and attending LTCF staff.

**Table 1:** Overview of core themes including description of theme and sub-themes.⍰.

| Theme name/subtheme | Definition of theme |
| --- | --- |
| 1. A shared commitment to providing care to residents. | Staff members associating their role with a duty of care to residents, which, at times, was stronger than the potential risks of the COVID-19 pandemic to the individual. |
| 2. Strengthening working relationships between staff members | Staff members felt close to one another due to increased time working together under challenging conditions. |
| a) Creating bonds through shared experiences | The shared experience of working in an LTCF during the pandemic facilitated bonding between staff members, subsequently improving teamwork. |
| b) Developing a team ethos | The development of a culture of reflection and shared practices to manage the ongoing challenges associated with the pandemic. |
| 3. Adapting to novel and changing roles | Staff adapting to duties and roles which were outside the scope of their original duties or job specification, in response to LTCF needs. |
| 4. Working as an incentive for socialisation | Staff actively choosing to continue working in the LTCF for the opportunity to socialise with other staff members and residents. |
| 5. Leadership by example | The behaviours of formal and informal senior staff members as a source of role modelling, emotional support and morale for junior staff. |
| 6. Recruitment and retraining of new staff | Difficulties experienced related to staff recruitment, retention and training of newly recruited staff members from outside of the long-term care sector. |
| 7. Divisions between furloughed and attending LTCF staff | Conflicted feelings towards staff members who chose to furlough or shield and their return to the facility. |

### 1. A shared commitment to providing care to residents

The most prominent theme emerging from the data was the feeling among staff that, by virtue of their role as working in an LTCF, they had a duty of care towards their residents. This duty of care, and the responsibility staff members felt towards the residents in their care surpassed any uncertainty or fear around the potential risk of COVID-19 infection to themselves. For some, a duty of care was an expectation of being a carer but also of other members of staff working in the profession. Within this belief, there was a central responsibility to keep attending the workplace, despite the risks of catching COVID-19 and being compensated with comparatively low pay compared to other key workers.

> *‘You take up a caring role, you know you’re going to come across things, not a pandemic, but every day there could be an infection in the home, and who knows whether it would kill you if you caught it anyway, you don’t know, do you? So that there is, you know, a duty of care as well within your contract*.*’*

P002

> *‘I felt like I didn’t want to take it home to my children, but I didn’t want to leave my colleagues. I didn’t wanna leave these (the residents) because who was gonna look after these (the residents) if we just said, you know what, yeah, we’ll work from home, we aren’t staying. We had sort of like a duty to stay because they’re not… their residents on paper, aren’t they? But to us, we know them and their families, and we can’t just leave them*.*’*

P008

At times, staff members felt conflicted between the choice to stay at home to protect their own families and attending work to provide care for residents who they also viewed as part of their family. During periods when the contact residents had with their own families was limited, such as during visitation restrictions, staff felt a responsibility to residents to be ‘their family’, in addition to providing care in a formal role. The commitment to caring for residents was a constant responsibility that extended beyond working hours and time spent within the facility. Some staff members reported either not engaging in or restricting the social activities they took part in outside of working hours to reduce the risk of infecting residents on their return to their facility.

> *‘It was like a family… it was a really nice time and I know I shouldn’t say that, but they were like a family, they (staff) were looking after the residents and not putting them at risk, they were concerned and they weren’t doing things outside of work because they didn’t want to put the residents at risk, which was, it was really good, it was a really bonding time I think*.*’*

P014

> *‘Because they (residents) don’t have, some of them, don’t have families and that, they are like family, they mean a lot to us, even when, I say we’ve got somebody poorly the other week, and I was messaging the girls, how are they? Are they OK? Even when I’m at home, so you don’t switch (off) …, they (staff) don’t switch off when they go home, they are still thinking about them and when it was COVID-19, that was even more ‘Oh, such-a-bodies got COVID-19, are they OK? Are they alright, please tell me they’re OK, oh thank goodness for that*.*’’*

P020

The duty of care meant that it was difficult for some staff members to ‘switch off’ at the end of their shift, with some checking up on residents after they had returned home or were outside the facility. Being a carer became part of the staff members identity, as opposed to a role undertaken on entry to the workplace, blurring the boundaries between the workplace and the home.

### 2. Strengthening working relationships between staff members

The increased workload that emerged during COVID-19, in terms of spending longer hours at the facility and working with a reduced number of staff facilitated the creation of closer relationships between staff members. Two subthemes emerged within this theme; creating bonds through shared experiences and developing a team ethos.

#### a) Creating bonds through shared experiences

The shared experiences of working in the LTCF during the pandemic facilitated the development of new bonds and strengthening of existing bonds between staff members. The increased opportunities for bonding were credited with improving teamworking, making the team ‘stronger’ and meant that staff were ‘there for each other’ in ways which they hadn’t been prior to the pandemic. For many LTCFs, the initial weeks of the pandemic were characterised by a reduction in the number of staff members working in the facility, due to a combination of staff isolating with suspected COVID-19 infection, staff shielding as either they themselves or someone in their household was vulnerable to infection, staff being furloughed, or staff simply refusing to attend work out of fear. The staff members that did remain in the facility were therefore fewer, working longer hours and spending prolonged periods of time working with the same co-workers. This resulted in stronger bonds between these workers, which were a source of support for staff members during periods of difficult working conditions.

> *‘They were difficult times, but in in some ways I think it brought the team quite close because it’s like a shared experience, isn’t it? And you know, it’s like the spirit of the blitz*.*’*

P019

> *‘I mean, the only good thing to come out of it was that the people who are still here, it brought us closer together. We are a very close-knit team*.*’*

P013

> *‘If you’ve got a problem, well, we can always help one another and everything, we got through it*.*’*

P024

In addition, staff bonding was facilitated by working in smaller teams due to the high numbers of staff shielding, or in cases where teams were restricted to working in specific area, i.e., on certain floors or areas of the building with extra infection prevention and control (IPC) measures. The working relationships developed during the pandemic created bonds which also extended beyond time in the facility.

> *‘It was difficult at the time, but it was good at the same time*… *because all the staff supported each other, we became a stronger team. Yeah, we’ve even got a little group chat so we can talk to each other to make sure we’re all OK*.*’*

P023

> *‘I think it brought the staff together as well… one of the girls, she had COVID, and we call her the COVID isolation buddies, (we) used to text each other all time or ring each other ‘what you’re doing? Yeah, just in your room, watching telly, been banned to my room*.*’*

P020

These working conditions also provided opportunities for compassion and shared support within teams which would not have been experienced under normal conditions. Staff began contacting each other outside of working hours to a greater extent than prior to the pandemic and supported each other with shopping or care packages if a staff member was isolating at home.

#### b) Developing a team ethos

Managing the potential spread of COVID-19 within the facility also required new ways of approaching the delivery of care. One benefit that emerged was the culture of ongoing learning that became established in some of the LTCFs interviewed, whereby staff reflected and shared their own practices with wider team members to solve problems. This could mean working together as a team to suggest new ideas, some of which were more stringent than the guidance issued by the government. In addition, LTCFs became more flexible in their approach to working practices, for example, changing the timings of shift work to suit staff needs.

> *‘It was a complete team effort and probably, if you were to look at it, just from that ethos that, team ethos, I think we were probably the best we were at that time because everyone was united in wanting to make sure we got it right*.*’*

P018

> *‘You know, one person said ‘well if we can use this room, we could set this up and do this, that and the other’, and I said, well, I’ll order more red bags so that we can put clothes in to take home without them contaminating anything else, you put them straight in your washer, all sorts of things like where all the ideas came in from the staff together. We worked together and that was what we wanted, it was probably more than what the guidelines were, but to us, it made us feel safer to do it that way*.*’*

P015

The risk of COVID-19 transmission was attributed to creating a sense of shared goals and values within the staff team. There was a mutual understanding on what the role of the staff members who were attending work was, and why they were doing it. This unified the team, and created a mentality whereby it was felt that all staff members took on equal or additional shares of the duties with little conflict or resentment. This mentality created a less hierarchical dynamic within the teams, with everyone listened to regardless of their background or job description, allowing staff voices to be heard.

> *‘It’s more like nobody was complaining of anything, … nowadays, it’s more like “oh, he’s not doing the job, he is also getting paid what I’m getting paid” but then it wasn’t like that. It was more nobody complains about it… It’s just that finding the staff was difficult and also the staff were like, nowadays it’s more like people will say “it’s not my job”, some of the things then it wasn’t like that during COVID, even with the four staff here, they will be like “OK, come on, let’s get on with this, let’s do this, let’s do that”*.*’*

P009

### 3. Adapting to novel and changing roles

The challenges presented by COVID-19 pandemic required staff members to adapt to changing roles, which were at times out of the scope of their original duties or job specification. In doing so, this meant staff members were working with other wider team members across the LTCF whom they previously would not have interacted or worked with on a regular basis.

> *‘We were running on empty, on minimal staff and I’ll be honest, how we did it I don’t even know, I don’t even know how we did what we did. You know, the cooks were cooking, coming down helping, it was just ridiculous, honestly*.*’*

P011

> *‘I think the staff became more supportive of each other and maybe a little bit more…. I mean within the teams, you have your sort of your housekeeping team, your kitchen team, don’t you, your care team, you tend get on to do their own bits, but in that scenario we all bonded together more and I think it became more teamwork rather than department work, if you know what I mean, so people supported each other more in that way*.*’*

P002

In some cases, this involved staff members whose previous role did not include direct face to face resident care taking on additional caring responsibilities due to staff shortages. Some of these roles carried additional risks, such as those related to direct patient contact with residents suspected of having COVID-19, and negotiations between staff on how to manage the workload, and associated risks, were common. For example, for staff who felt uncomfortable with direct patient contact, some were offered administrative tasks or general housekeeping roles to ease the workload on other members of the team.

> *‘I wouldn’t force staff to go in (the room), I was just happy to have them in even if they were just taking meals out and things like that, making sure they (residents) were drinking, or happy to go in and do personal care*.*’*

P011

> *‘I don’t think anybody here is like, well, “you’re the cleaner and I’m the…” there’s nothing like that here really, we all see each other as an equal in one way, so I think just supporting each other really*.*’*

P008

Subsequently, the divides and hierarchies that previously existed between staff groups were reduced, contributing to weakening the hierarchy within the team. The boundaries of staff roles becoming less defined also had an impact on cooperation between staff, with fewer divisions in terms of job role and associated pay.

### 4. Working as an incentive for socialisation

At times of national lockdown, coming into work provided an opportunity for socialisation that was not available elsewhere or to the wider public. This was greatly appreciated by LTCF staff members and provided an incentive to take on additional shifts despite the potential risks.

> *‘I thought it was nice as well to see people, that you could actually be around other people. I mean, it must have been hard… for people at home all the time. … so, it was nice for us to be able to come and have different conversations*.*’*

P020

> *‘It was rubbish having to come to work because of the conditions that we had to work in, but at the same time, we were like, well, at least we get to come to work, we’re not stuck at home all day and staring at the clock and wondering what to do and wondering when to go out for our twenty minute walk*.*’*

P008

One of the main drivers for remaining in the LTCF workforce, despite the challenges and potential risks, was the opportunity for emotional support and camaraderie. Attending the facility each day allowed for social contact with other staff and residents, at a time when there were relatively fewer other opportunities for socialisation. This was especially so for older staff members and those who lived alone.

> *‘During the first (lockdown), people being off, not with COVID, but just being off in the first lockdown was fine because nobody could go anywhere, so everybody was quite happy to work as many hours as possible because otherwise they we were just sat at home with the other half or on their own or whatever*.*’*

P021

Some staff described the experience as like living in a bubble, where despite the challenges that were in the LTCF, it was a safe space and provided comfort in the face of wider uncertainty. It also provided staff with something to do, rather than be at home.

### 5. Leadership by example

One aspect of changing team dynamics during the pandemic described by staff members related to junior team members looking to senior staff for role modelling, emotional support and maintaining morale. In some cases, senior staff referred to managers or those in an official leadership role, in others it related to senior carers, those who were older or those who had worked in LTCFs for a relatively long time.

> *‘Even as a carer, I was always a leader. I was always like right we are doing this, probably a bit controlling, but I do believe that your staff will respect if you get on the floor and help them and do it instead of telling them, it’s worked for me for seven years. So yeah, I do support my staff because I know what it’s like, I know what it’s like to be a carer, I know what the job is*… *I do know because I’ve experienced all of them feelings. You know, when they feel like they’re not doing the job well enough*.*’*

P011

In another LTCF, the manager was well regarded for taking on a resident facing role and wearing a uniform, rather than staying in the office. Staff members who took on an active role in providing resident care were held in high esteem in general, due to choosing to accept the additional risk it posed to staff members and the benefits such care made to residents.

> *‘I don’t think I would have been able to get through it without the managers, I think if it would have been just carers. I don’t know if it’s because they have more insight of the medical side of it, so even though it was chaos you felt sort of safe within that bubble, that you were protected as long as everybody stayed within that bubble*.*’*

P013

### 6. Recruitment and retraining of new staff

One of the main challenges for LTCFs throughout the pandemic was a lack of available staff. Prior to the pandemic, adult social care was characterised by difficulties in staff recruitment and retention, pressures which were compounded by the pandemic. Initially, staff shortages were due mainly to staff either shielding, on furlough, isolating, or staff choosing to leave the profession due to fears regarding COVID-19 transmission. In one facility, the initial praise and recognition provided to care workers was attributed with encouraging more people to move into the sector, which benefited the facility.

> *‘We did employ more staff. I think staffing for health and social care is really, really difficult at the moment. I think there’s no*… *during the first lockdown, there was lots of praise and recognition for what people did, and I think people all think that’s very, very quickly been forgotten and getting good staff is really difficult, so I think if we had one again I think it would be even more difficult to recruit people as we did because I say we did recruit staff which worked really well, if we hadn’t done that, I think we would have struggled even more*.*’*

P021

However, in other facilities, the impact of staff shortages varied across the course of the pandemic. At times, existing staff members were keen to take on more shifts to reduce the financial impact of the pandemic on household income, reducing the need to bring in newly recruited or agency staff. In others, staff shortages were much higher and had greater implications for resident care. In relation to the government recruitment campaign to get new staff members into LTCFs, staff reported relatively few new starters. Those that did start working in the LTCF were reported as finding it difficult without prior training, and there was a sense that the general public did not fully understand what it was to be a care worker, in terms of the emotional support and physical endurance involved in a caring role.

> *‘They don’t have job now, so they can come to work as a carer, they were working somewhere else when they don’t have job. There is only sector which was running was the health sector, so they came to work on this… but whoever hasn’t worked in care, they only know to feed people, maybe even washing they struggled*.*’*

P009

### 7. Divisions between furloughed and attending LTCF staff

The data identified conflicting experiences and needs reported by those who remained working in the LTCF when reflecting on staff members who shielded, were furloughed or chose to remain at home. Interview participants were staff members who remained actively working in the facilities, and the topic of those who had chosen not to was discussed frequently. The staff who attended work were described as the strongest of those ‘left behind’, and the unification that occurred in this group resulted in low staff turnover, which was perceived as a sign of good teamworking. This mentality, while positive for those within the group, understandably created a divide, with those who did attend work finding it difficult to empathise with those who were either furloughed or actively chose not to attend.

> *‘I had one girl who went off because she was frightened, and she’s got two kids and a one parent family. I was not impressed because we’ve all got kids and quite a few members of staff didn’t have families and were one parent families, but she just said she wasn’t coming in and basically there was nothing I could do about it, but as I said to her, we’re all frightened, we’ve all got kids. What if every member of my staff had said that to me?’*

P002

> *‘I think that caused a bit of… I wouldn’t say a rift or anything, but I just think the ones that didn’t have the underlying health condition that left, I think well we all had a reason not to be here, but some of us stayed to support our colleagues, and our residents. I had children…. I didn’t want to be here but I couldn’t leave. I couldn’t just leave and leave everybody to carry on. And what would I do then? Just sit at home and wonder? I just couldn’t do it personally, so I think some of us felt like we’d been a bit let down by some of our own staff because we all had a reason not to work*.*’*

P008

The reasoning behind this divide was rooted in multiple perspectives, primarily related to the availability of furlough schemes, justification of risk and feeling let down. At times, staff members felt it was unfair that others were given the option for furlough, for both those working in the care sector, but also the general population. Others felt that the justification for not attending work, and the approach to risk by such workers, did not outweigh the duty of care that was expected of workers in this long-term care sector. This was especially so when participants with similar health conditions to those of workers who chose to shield themselves remained attending work. On an emotional level, there was also a sense of feeling let down by colleagues who chose to protect their families rather than attend work, given the resulting increase in workload for those who remained.

> *‘When the furlough had stopped and everything and they said about people coming, there was a bit of a thing between the staff as well because not only are they being furloughed and they want to all their holidays… so there was a bit of*…*they had to come back into the home gradually because people had… we’ve been through it, and we’ve all worked it, you’re taking advantage of it… it was all set up wrong*.*’*

P006

The return of furloughed staff after the pandemic increased the sense of unfairness and further deepened the divide. Animosity remained, with some staff feeling that those returning had avoided, and indirectly added to, the challenges remaining staff had experienced during the pandemic, and therefore should not be able to return to their job. In some LTCFs, managers chose to not allow furloughed staff to return to their old jobs and incorporated questions regarding willingness to work during pandemics into the recruitment process for newly appointed staff.

*Participant: ‘I can’t believe they run off. I can’t believe it. Ran off, cause these some of these people I’ve worked with for years and I just couldn’t believe*.*’*

*Interviewer: ‘Do you know if they left the profession entirely, or were they just off for a bit? Did they come back?’*

*Participant: ‘No, I think 90% of them left. Quite a few of them tried to return much later, so they’re still in the profession, but I wouldn’t have them back*.*’*

P005

## Discussion

This paper explored the experiences of staff working in LTCFs in England during the COVID-19 pandemic, using data collected in twenty-four semi-structured, qualitative interviews. In doing so, seven themes emerged, relating to a duty of care to residents, bonding through shared experiences, changing and adapting roles, incentives for socialisation, leading by example, and difficulties in recruiting, retraining and the acceptance of returning staff.

The findings of this paper support those of other studies which have explored the experiences of staff members in similar settings during the COVID-19 pandemic. In Canada, long-term care staff reported renewed feelings of passion for their role, the development of a close community among LTCF staff and a sense of pride in the novel and creative approaches that they developed in response the challenges faced (22). In a similar European study, participants stated that a good atmosphere existed within the team during the pandemic, and that consistent teamworking with the same co-workers strengthened cohesion by providing opportunities for staff members to talk to each other, facilitating mutual support and encouragement (23). LTCF managers viewed the skills and determination shown by staff as a source of resilience and demonstrated creativity and skills that had previously not been required or utilised (24). In addition, staff members have reflected positively on the affect that the pandemic had on their relationships with their wider team, and the benefits strengthening these bonds had on their own individual empowerment and resilience, specifically in terms of supporting staff members during times of additional pressures and uncertain risk.

The findings also demonstrate the flexibility and willingness of staff members in terms of their ability to adapt to change. This was especially so for staff members that took on new or additional roles to provide direct personal care to residents, such as staff previously employed in roles others than carers, including housekeepers or catering (25). Again, these experiences are not unique to the UK long-term care sector - in one US based study, 68% of participants reported performing tasks that were outside of the original scope of role, 62% reported an increase in time required to complete tasks, and 27% reported added pressures in their workload (26). In this study, LTCF staff reported that the pressures and challenges associated with the pandemic provided an environment in which both communication and teamworking skills could not only develop, but flourish in ways which hadn’t been in place prior. These two subthemes of strengthening working relationships between staff members identified; creating bonds through shared experiences and developing a team ethos, are not mutually exclusive, and frequently reinforced one another. The findings of this paper show that the pandemic inadvertently created conditions which facilitated improved team working and working relationships between some staff members, most notably those who continued to work in the facility.

The findings also raise questions regarding the potential mediating effect of staff working conditions on mental health. Satisfaction with the management of COVID-19 by the facility manager, support at work and a sense of community have been associated with clinically lower levels of stress, anxiety, and depression among the LTCF workforce (27). In a survey of frontline health and social care workers in the UK, 58% met the criteria for a clinically significant disorder, such as anxiety, depression, or post-traumatic stress disorder, and those who felt that they could not talk with their managers if they were not coping or who reported feeling stigmatised were more likely to meet criteria (28). In an interventional survey comparing staff experiences of stress and anxiety in Sweden, Italy, Germany, and the UK, not only were total stress and anxiety levels highest in the UK, but having support from management was associated with lower stress and anxiety levels (29). The benefits of improved teamworking are not only likely to improve the quality of care provided to residents, but also have the potential to act as a protective factor in mediating the effect of a pandemic on mental health and wellbeing.

However, despite the positive implications on staff working relationships, these findings are very much within the context of a wide literature illustrating the negative impacts of a pandemic on staff mental and emotional wellbeing, specifically in terms of increased rates of anxiety and depression, across LTCFs internationally (30-32). In this study, the focus of the analysis has been on the working relationships between staff members, however participants also reported the negative impacts of the pandemic on their mental and emotional health and wellbeing, which have been reported in detail elsewhere (33). These co-existing experiences have been identified in other studies, which showed that while LTCF staff described higher levels of work-related stress and burnout, staff still conveyed a sense of pride in their role of caring for residents (34).

### Strengths and limitations

A strength of this paper is its focus on working relationships, as opposed to staff experiences in general. Participants were initially asked to share their experiences of working in LTCFs during the pandemic, specifically on implementing the policies issued by the UK government, and the barriers and facilitators to implementing these policies. This meant that, outside of prompts regarding the policies issued, participants had the freedom and space to discuss their experiences, focusing on the topics which they felt strongly about. Subsequently, thematic analysis directed the secondary publications from the original study, so the themes related to staff working relationships emerged organically from the data.

The generalisability of the findings is potentially limited compared to other studies, due to recruitment being restricted to one region in England. However, this allowed LTCFs who were subject to similar local restrictions to share their experiences, improving comparability across the interviews. The approach to recruitment also excluded facilities which were rated as inadequate by the Care Quality Commission, therefore the staff working relationships in these facilities were not captured in this study. In the context of the pandemic, it is possible that the positive experiences discussed in this paper were an artifact of LTCFs which were performing relatively well prior to the pandemic. Similarly, LTCF staff who had subsequently left the facility and not returned were not recruited to the study, so these experiences are also missing from the narrative.

The retrospective approach to data collection has both advantages and disadvantages which require further discussion. In conducting the interviews one to two years after the final restrictions were imposed, LTCF staff were also able to reflect on their experiences with colleagues who had previously been furloughed or shielded at home returning to the workplace. The divisions between these two staff groups and the influence this has had on recruitment practices are important findings from this study that would have remained unexplored had this research been conducted during the pandemic. However, there is a risk of recall bias, and the potential for staff viewing their experiences through ‘rose tinted glasses’, whereby the past is recalled more positively than it was experienced at the time.

### Implications for research, policy and practice

The findings provide an insight into the experiences of LTCF staff working during the pandemic, primarily that LTCF staff reported a sense of duty at a time of uncertainty. Despite the personal risks staff were taking in attending work, those who remained were able to form strong support networks, which allowed them to continue to provide care to residents. Further research is needed to understand how this approach to teamworking can be created in less challenging conditions, and the facilitators and barriers which support this. In addition, the relative importance of improving staff teamworking within the context of wider strategies previously identified as integral to improving quality of care, such as providing leadership, training opportunities, alongside better pay and working conditions, requires further thought (35).

In addition, the findings highlight two key practices which could further prepare LTCFs for future pandemics. Firstly, providing training across roles so that direct resident care can be delivered by all staff members in times of limited staffing. The success of decreasing the hierarchy between different staff groups, working across teams and staff members taking on novel or additional roles has been discussed at length by the participants in this study. Secondly, the positive effects discussed are within the context of difficulties in recruiting and retaining staff within the long-term care sector, which have been exacerbated by the pandemic. Staff vacancy rates and retention rates have been found to correlated with the likelihood of being rated as ‘Good’ or ‘Outstanding’, at a time when the number of filled positions in the adult social care sector in 2022/23 was still below its pre-pandemic peak in 2020/2021 (by around 45,000 filled posts) (13, 36).

In applying a policy perspective, a focus should be on how to manage the reintegration of staff who have returned from furlough, and how these staff members can be successfully incorporated back into the LTCF. At a time of staff shortages within the sector, retaining experienced staff is a priority. There are also implications for recruitment of new staff and establishing the willingness of potential staff to continue working during pandemics. The legality of this practice is complex, however further guidance is needed to establish fair and equitable approaches to the risk assessment of potentially vulnerable staff members, and how this can be managed in a way that allows a facility to maintain staffing ratios during pandemics without putting additional workloads on remaining staff members.

Finally, the findings of this study have shown how much of the benefits discussed were underpinned by a shared sense of duty to the residents within the LTCF. The experience of working within an LTCF is often characterised by low pay, limited opportunities for progression and dealing with difficult situations associated with caring for older adults with dementia or at end of life. The participants in this study showed commitment and loyalty to the long-term care sector despite these working conditions. An understanding of how staff members can be appropriately compensated will undoubtably support both recruitment and retention of this workforce.

## Conclusion

LTCFs experienced substantial challenges during the COVID-19 pandemic, however within these conditions staff reported developing a strong level of teamwork among those who remained working within the facility. These bonds were characterised by a shared experience of adapting to new roles, working within smaller teams, and looking to more senior staff for leadership. External to these teams, reintegrating staff members who left the facility, and introducing staff recruited from outside the social care sector were areas LTCF staff felt conflicted about. Further research is needed on how to support LTCF staff to develop and strengthen their teams prior to future pandemics, and how the positive impacts of the COVID-19 pandemic on staff working relations can be sustained.

## Data Availability

Data is available on reasonable request from the authors.

## Data Availability

Data is available on reasonable request from the authors.

## Abbreviations

CHAP: Care Home Assistant Practitioner
IPC: Infection prevention and control
LTCF: Long-term care facility

## Ethics approval and consent to participate

Ethical approval was obtained from the Lancaster University Faculty of Health and Medicine Research Ethics Committee (reference: FHM-2023-3368-RECR-3)

## Consent for publication

Written informed consent was collected from both the manager of the LTCF recruited to the study and the LTCF staff member participating in the interview for anonymised publication of the data collected, including quotes.

## Data availability

Data is available on reasonable request from the authors.

## Clinical trial number

Not applicable.

## Authors’ contributions

DCM conceived the study, developed the study protocol, applied for ethical approval, and prepared the manuscript. NC coordinated recruitment, conducted the interviews and contributed to the overall writing of the paper. DCM and NC analysed the data.

## Acknowledgments

The authors would like to acknowledge and thank the Enabling Research in Care Homes network for supporting the recruitment for the study from which this data was collected. The authors would also like to thank the LTCFs and staff members who took part in this research for their valuable contribution.

